# Clinical burden of disease and demographics in older adults hospitalised with RSV infection in England: A retrospective cohort study

**DOI:** 10.64898/2026.07.21.26358579

**Authors:** Rebecca Butfield, Kiran K. Rai, Thomas Jennison, Jack Said, Hannah Wright, Dheeraj Sethi, Jonathan Watkins, Antonia Geneidat, Isabel Jimenez, Dexter Wiseman

## Abstract

**Introduction:** Respiratory syncytial virus (RSV) causes significant disease in older and comorbid adults. Current UK vaccination recommendations restrict eligibility to adults ≥75-years, 65-74-years with chronic respiratory disease or immunosuppression, and those in care homes, but evidence on clinical burden in adults <75-years with comorbidities is limited. This study assessed patient characteristics, healthcare resource utilisation (HCRU), and mortality in adults hospitalised with RSV in England.

**Methods:** Population-based retrospective cohort study using linked Clinical Practice Research Datalink Aurum and Hospital Episode Statistics. Adults ≥60-years hospitalised with RSV between October 2014-March 2019 were included. RSV episodes defined as 90-days post diagnosis. Case definitions were created using diagnosis codes related to confirmed RSV (RSV-specific) or acute lower respiratory tract infection where other causative pathogens were excluded (RSV-possible). All-cause HCRU (hospitalisations, critical care admissions, outpatient attendance, primary care consultations, and prescriptions) and case fatality rates were assessed. Results were stratified by age (60-74 and ≥75-years), case definitions (RSV-specific, RSV-possible) and comorbidity profiles (chronic respiratory disease, immunocompromised, cardiovascular disease).

**Results:** A total of 97,712 hospitalised episodes in those aged ≥60-years were included in the analysis, where 785 (0.8%) were RSV-specific cases (n=338 aged 60-74-years, n=447 aged ≥75-years). In RSV-specific cases, median (IQR) cumulative length of stay (LoS) for those ≥75-years was 10.00 (6.00-22.00) days which was equivalent to or lower than each comorbidity sub-group in those aged 60-74-years, with longest LoS in those immunocompromised (11.50 [7.00-26.00] days). Critical care admissions were more often observed across comorbidity stratifications (13.85%-22.34%) compared to those ≥75-years (5.03%). All-cause and RSV-related case fatality rates for RSV-specific cases were highest among those ≥75-years (all-cause: 19.3%; RSV-related: 10.3%).

**Conclusion:** HCRU within RSV episodes in those aged 60-74-years across comorbidity profiles was equal to or greater compared to those aged ≥75-years. These findings highlight the need to prioritise consideration of comorbidity groups that could benefit from RSV vaccination.

**Key summary points:** *Why carry out this study?:* • There is growing evidence on the burden of disease of RSV, however there is still a question around whether everyone at risk of severe outcomes are covered by the UK vaccination programme. • There is a paucity in evidence on the clinical burden of RSV in adults <75-years, especially in those with comorbidities including cardiovascular risk factors. Granular HCRU and morality outcomes are required to inform policy decisions on RSV vaccination strategies in these groups. • This study aimed to quantify the clinical burden of RSV in adults ≥60-years in England, stratified by comorbidity profile and descriptively compare to those previously eligible for the RSV-vaccine programme in England (aged ≥75-years, and in those in adult care homes).

*What was learned from the study?:* • The study found that the clinical burden of RSV in secondary care in adults aged 60-74-years across all comorbidities stratifications, is equivalent to or exceeds the burden of those aged ≥75-years.

## Introduction

Respiratory syncytial virus (RSV) is one of the leading causes of acute respiratory tract infections within adults, causing upper and lower respiratory tract disease (URTD, LRTD) (1, 2). Infection can progress to severe disease, often associated with exacerbation of preexisting comorbidities, increased risk of hospital and intensive care admissions, and mortality. Populations at high risk of severe infection include older adults, and those immunocompromised or with chronic conditions (2–4).

In the United Kingdom (UK), RSV infections follow a seasonal pattern, typically peaking between October and March (5). Whilst recent estimates (Winter season 2023/24) suggest RSV-associated hospitalisations (58.3 admissions per 100,000 people) tend to be lower compared to other respiratory viruses (e.g. 114.6 per 100,000 people for influenza-associated hospitalisations) (6), diagnostic testing for RSV is infrequently performed in adult patients (4). Lack of universal testing is partly due to suboptimal sensitivity of available tests. In a recent meta-analysis, pooled sensitivity for identifying RSV using rapid antigen detection and direct fluorescent antibody testing in comparison to multiplex polymerase chain reaction (PCR) testing was 64% (95% CI, 50%–75%) and 83% (95% CI, 66%–93%), respectively (7). However, using multiple diagnostic methods can increase RSV diagnostic yield by 2.6-fold (8). Furthermore, there is a lack of RSV-specific treatment where severe cases are often managed through supplemental oxygen and mechanical ventilation (6) meaning a specific RSV diagnosis is not required for the delivery of standard care. Where the presence of RSV is confirmed, RSV-specific ICD-10 coding is rarely observed in the patient record (6%), however when combined with related codes indicative of acute lower respiratory infection, 44% of RSV confirmed cases were identified (9). Subsequently, the true disease burden for RSV in adults is likely underestimated.

Three RSV vaccines are currently approved for adults by the UK Medicines and Healthcare products Regulatory Agency (MHRA) and the European Medicines Agency (EMA). In September 2024, the UK Joint Committee on Vaccination and Immunisation (JCVI) recommended adults aged 75-79-years to be offered RSV vaccination (10). In March 2026, the committee expanded the recommendation to include those 80-years and above and residents in older adult care homes (11), with those 65-74-years of age with chronic respiratory disease or immunosuppressed made eligible in April 2026 (12). The JCVI advised that considerations on extensions to other groups should be guided by emerging epidemiological evidence on RSV burden in these groups.

There are limited studies that quantify the clinical burden of RSV-infections in adults under the age of 75 in the UK, specifically those with comorbidities. A recent study that pooled UK hospital cohorts. demonstrated age-based heuristics showed a lower ability to predict the risk of ICU admission/mortality compared to heuristics that considered age and comorbidities in adults hospitalised with RSV (13). Previous literature has highlighted individuals with cardiovascular disease, chronic respiratory conditions and immunocompromised disease are at higher risk of developing severe RSV infection and related complications (14–16). Further research demonstrates a relationship between RSV and cardiovascular outcomes; in a systematic review and meta-analysis, pooled estimates across 25 studies demonstrated that following RSV infection, 19.2% of RSV patients experienced a cardiac event, and 15.7% and 5.4% experienced heart failure and acute coronary syndrome, respectively (17). Moreover, in a prospective cohort study of 377 patients with COPD, RSV was associated with 8.7% of outpatient-managed COPD exacerbations (18). In a further study, immunocompromised individuals have been shown to have worse outcomes when contracting RSV, including prolonged length of stay in hospital and death (19, 20).

Despite previous research reporting a heightened risk of severe RSV-related outcomes among adults with specific comorbidities, there is limited evidence on the granular clinical burden, such as healthcare resource use, in these groups. Therefore, this study aims to assess the patient characteristics, health care resource utilisation and mortality in adults hospitalised with RSV in England, stratified by comorbidity profile, across a 90-day episode. By addressing this evidence gap, we will provide valuable evidence to support policy makers and clinicians in making informed decisions about RSV vaccination strategies.

## Methods

### Data source

A population-based retrospective cohort study was conducted using primary and secondary care data from England. Primary care data from the Clinical Practice Research Datalink (CPRD) Aurum database (December 2024 release) were linked to secondary care data from Hospital Episode Statistics Admitted Patient Care (HES APC), Outpatient (HES OP), the 2019 English Index of Multiple Deprivation (IMD) and the Office of National Statistics (ONS) datasets.

CPRD Aurum is a longitudinal, representative anonymised electronic health record (EHR) database of primary care interactions in the UK, covering ∼20% of UK general practices as of December 2024. There are over 35 million patients with data records eligible for linkage and approximately 16 million of whom are active (i.e. still alive and registered with the General Practitioner practice) (21). Data captured include patient demographics, medical diagnoses, care received in primary care, lifestyle information, and prescriptions issued in primary care.

HES APC and OP datasets contain details on inpatient admissions and outpatient appointments, respectively, to NHS funded centres in England. At the time of this data acquisition, the latest release of HES APC and OP data available through linkage to CPRD covered the periods April 1997 to March 2023 and April 2003 to March 2023, respectively (22, 23). IMD data is based on 39 indicators combined into seven weighted domains to provide a measure of socioeconomic status (24), and death registration data were available in the ONS database.

This study was approved by CPRD’s Research Data Governance (RDG) Committee (reference number: 24_004459).

### Study design and population

Adult patients aged ≥60-years in England hospitalised with an RSV diagnosis between 1^st^ October 2014 and 31^st^ March 2019 (indexing period) were included in the study. The study design was based on an episode-level analysis, where an RSV episode was defined as 90-days after an RSV diagnosis code (index date); as such, patients may have had multiple RSV episodes and were therefore represented as separate index events referred to as episodes through the rest of this manuscript (25, 26). The study design is illustrated in **Figure 1**. A RECORD reporting guidelines checklist is available within the **Supplementary Material**.

**Figure 1.**
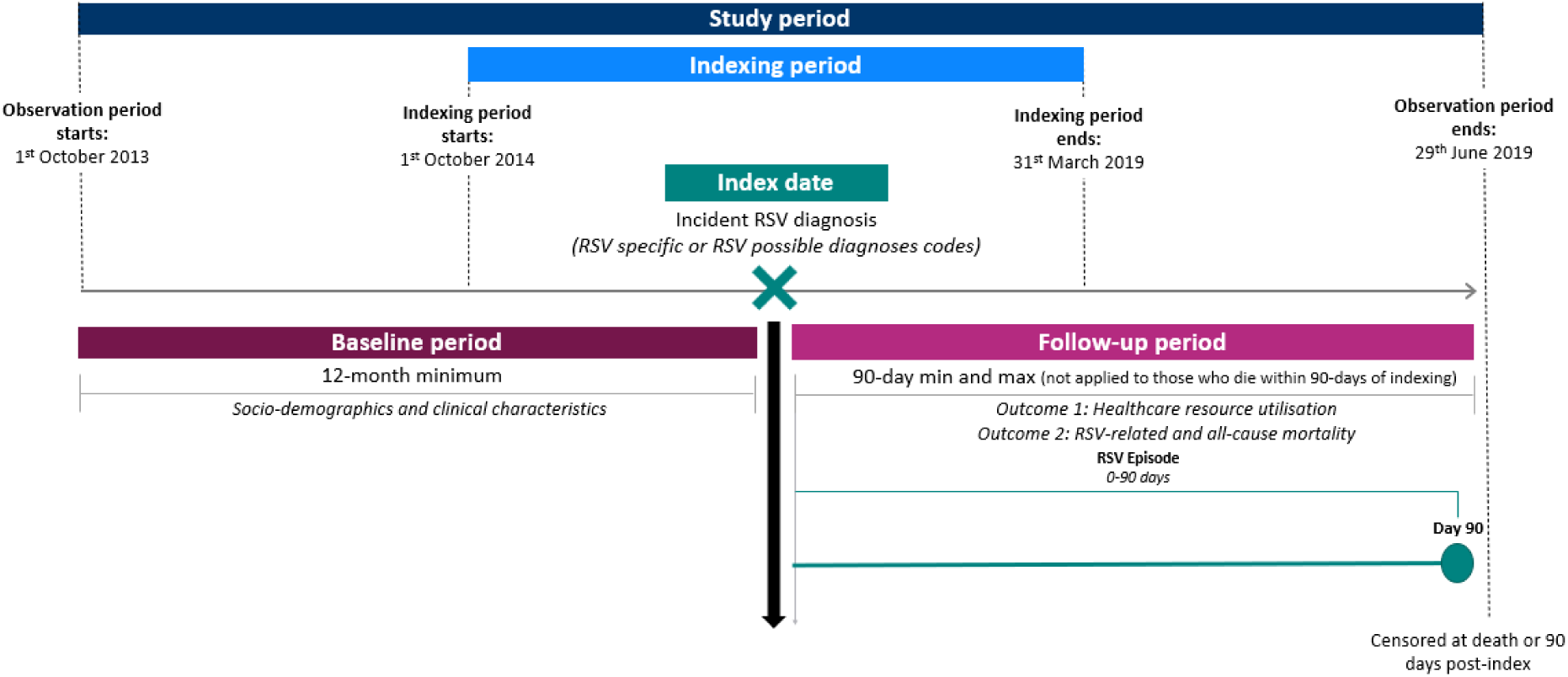
Study design schematic. An RSV episode was defined as 90-days after an RSV diagnosis code (index date); as such, patients may have had multiple RSV episodes. The indexing period was the time whereby hospital episodes with an RSV-specific or RSV-possible diagnosis code were observed for inclusion. The observation period was defined as 12-months prior to index, where baseline characteristics were assessed, and 90-days following index, where HCRU and mortality were assessed. Patients were censored at the earliest of: 90 days post-index or patient death.

**Figure 2.**
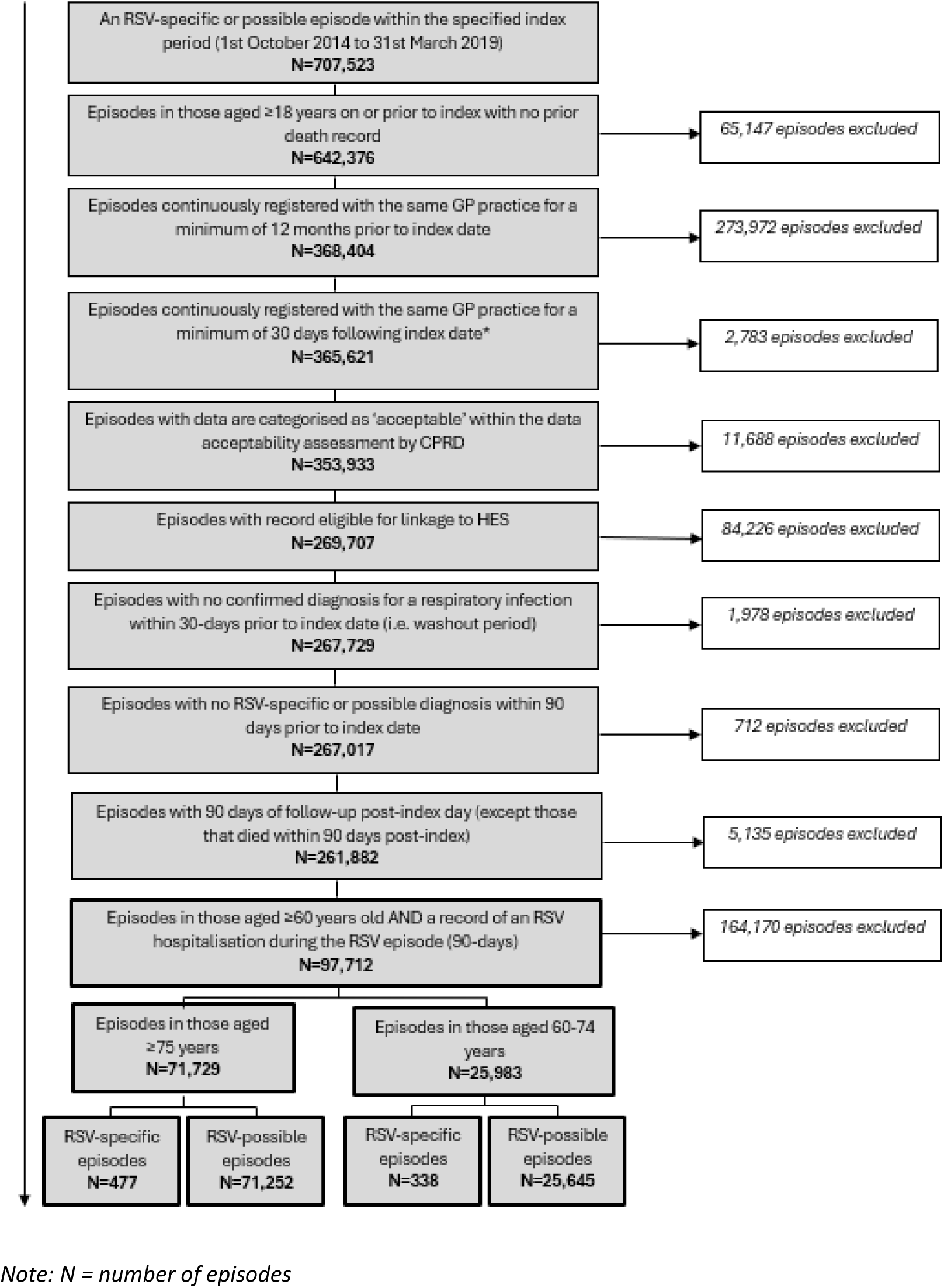
Episode attrition

Episodes with an RSV-specific or possible diagnosis in either the primary or secondary diagnosis position within secondary care (HES APC) were included. The index date for each episode was defined as either:

- The hospital admission date if no primary care RSV diagnosis code was recorded within 90 days prior to the hospitalisation
- The primary care diagnosis date if an RSV-related hospitalisation had a corresponding RSV-specific or possible diagnosis recorded in primary care (CPRD Aurum) within 90 days of and including the hospitalisation admission date.

Episodes were also required to meet the following criteria to be eligible for inclusion: aged ≥60-years at episode index date; general practitioner (GP) practice registration ≥12 months prior to and ≥90 days post-index (except where a death was recorded within the first 90 days post-index); primary care record eligible for linkage with HES; and data considered of acceptable research quality as defined by CPRD (21). Episodes were excluded if a respiratory infection was observed within 30-days prior to the index date (see **Supplementary Material** for excluded respiratory infections).

### Exposures

RSV diagnoses were identified using diagnosis codes recorded via International Classification of Diseases and Related Health Problems, 10^th^ Revision [ICD-10] codes or SNOMED codes, within secondary care non-elective inpatient admissions and primary care, respectively. Two distinct case definitions were created based on the presence of specific and possible RSV diagnosis codes (9). The RSV-specific case definition consisted of RSV episodes where observed diagnosis codes were indicative of RSV-infection. The RSV-possible case definition consisted of RSV episodes where observed diagnostic codes were indicative of generalised acute lower respiratory infection and would therefore capture episodes that are possible to be caused by RSV. Episodes that were indexed based on RSV-possible diagnosis code(s) but then had an RSV-specific diagnosis code observed within 14 days of index were reported as an RSV-specific episode.

Code lists were adapted from published literature, reviewed by the study team and verified by those with clinical experience within the UK health service (see **Supplementary material** for code lists) (27, 28).

### Demographic and clinical characteristics

Sociodemographic characteristics at index included: age; sex; ethnicity (using entire medical history); socioeconomic status (measured using quintiles of the 2019 IMD score based on the patient’s postcode); and care home residence status within 5-years prior to index. Clinical characteristics included: smoking status (using entire medical history); body mass index (BMI) within 5-years prior to index; comorbidities of interest (using entire patient history); Quan-Charlson Comorbidity Index 2011 (CCI) (29) using entire patient history; index year; and whether the index event occurred during a RSV season (defined as occurring between October and March, inclusive)(5).

### Health Care Resource Utilisation

#### Outcome assessment period

HCRU and case fatality rates were reported during the RSV episode, from index (day 0) to day 90 post-index. Patients were required to have complete follow-up (90-days) for each episode to be assessed within each period, unless they died prior to the end of follow-up. In cases where they die prior to 90-days, patients are assessed up until the point of death (i.e. censor date).

Health care resource utilisation (HCRU) for each RSV-episode (90-days post-index) were described for the following elements of HCRU:

#### Hospitalisations

All-cause general inpatient admissions and, the cumulative length of stay (LoS) in general inpatient per episode, were reported. Additionally, critical care admissions (high dependency/intensive care unit) and the cumulative LoS in critical care per episode were reported separately. The following critical care interventions were also reported: non-invasive ventilation or continuous positive airway pressure (CPAP); and invasive ventilation or extracorporeal membrane oxygenation (ECMO). All-cause hospital readmissions within 30 days of discharge from an RSV-related hospitalisation were also reported.

#### Outpatient specialist attendance

RSV-related outpatient specialist encounters observed per episode were reported, defined as a maximum of one attendance per-person per-day See **Supplementary material** for further information on specialities considered to be potentially RSV-related.

#### Primary care consultations

All-cause GP or nurse consultations were reported per episode. This was defined as a maximum of one visit per-person per-day; any additional visits were considered as data capture errors.

#### Primary care medication use

Medications that were prescribed within primary care during the RSV-episode were reported for the following: antibiotics, bronchodilators; diuretics; inhaled corticosteroids; and oral steroids.

#### Mortality

All-cause and RSV-related case fatality rates were calculated as the number of death events divided by the number of RSV cases during the 90-day episode. RSV-related fatalities were defined as death events with an RSV-specific or possible diagnosis ICD-10 code identified as the primary or secondary cause of death in the death certificate obtained from ONS.

## Statistical analysis

This study did not involve hypothesis testing; therefore, no formal sample size calculations were performed and data are described numerically. Means and standard deviation (SD), median and lower and upper quartiles (Q1, Q3), and minimum and maximum values were calculated for numerical variables, with frequency counts and percentages presented for categorical variables. HCRU outcomes were reported for resource users only. Results on <5 patients were suppressed to comply with CPRD reporting rules, with secondary suppression implemented where relevant.

Results were presented for RSV-specific and RSV-possible episodes occurring in those 60-74-years of age, and separately, for a cohort aged ≥75-years. Results for the 60-74-years of age cohort were also stratified into subgroups by comorbidity profiles associated with high risk of severe RSV infections (immunocompromised disease, cardiovascular disease [excluding essential hypertension] and chronic respiratory conditions (30). See **Supplementary material** for the list of comorbidities included in each subgroup.

## Results

In total, 97,712 hospitalised RSV-specific and RSV-possible episodes were identified in 89,138 patients, aged ≥60-years, occurring between 1^st^ October 2014 and 31^st^ March 2019 and were included in the study (**Figure 3**). Of these episodes, 25,983 (26.6%) were in those 60-74-years of age, with 338 episodes identified via the RSV-specific case definition and 25,645 via the RSV-possible case definition. A total of 71,729 (73.4%) episodes were identified in those ≥75-years of age; 477 of these episodes were included in the RSV-specific case definition and 71,252 via the RSV-possible case definition.

### Demographic and clinical characteristics

#### RSV-specific case definition

Of the 338 episodes (in 336 patients) aged 60-74-years, 60.4% (n=204) had cardiovascular disease, while 58.3% (n=197) and 19.2% (n=65) had chronic respiratory conditions and an immunocompromised disease, respectively **(Table 1** and **Table 2)**. The mean age was 67.4 (SD: 4.2) years and 55.6% were female. The majority were of White ethnicity (88.2% [n=298]) and roughly half of episodes were in the most deprived quintiles of socioeconomic status (quintile 4: 24.3% [n=82]; quintile 5: 22.2% [n=75]). Approximately 80% of episodes were either current or former smokers, and 17.4% (n=59) were classified as being either obese (BMI ≥30-<40 kg/m^2^) or severely obese (BMI ≥40 kg/m^2^). Over 90% of episodes were indexed within a respiratory season. The mean Quan-CCI was 2.5 (SD: 2.2) and median length of follow-up was 90.0 (Q1, Q3: 90.0, 90.0) days.

**Table 1.** Baseline demographics characteristics among **RSV-specific and RSV-possible** episodes in those ≥75-years and 60-74-years of age, stratified by comorbidity profiles.

| Demographics | $\geq 75$-years | | 60-74-years | | 60-74-years comorbidity profiles | | | | | |
| --- | --- | --- | --- | --- | --- | --- | --- | --- | --- | --- |
|  |  |  |  |  | Chronic respiratory condition |  | Immunocompromised disease |  | Cardiovascular disease |  |
|  | RSV-specific<br>(N=477) | RSV-possible<br>(N=71,252) | RSV-specific<br>(N=338) | RSV-possible<br>(N=25,645) | RSV-specific<br>(N=197) | RSV-possible<br>(N=13,490) | RSV-specific<br>(N=65) | RSV-possible<br>(N=1,277) | RSV-specific<br>(N=204) | RSV-possible<br>(N=16,876) |
| **Age at index (years), n (%)** |  |  |  |  |  |  |  |  |  |  |
| *Mean (SD)* | 84.10 (6.47) | 85.33 (6.13) | 67.44 (4.23) | 68.31 (4.17) | 67.46 (4.40) | 68.50 (4.11) | 67.26 (3.90) | 68.06 (4.14) | 67.79 (4.33) | 68.69 (4.07) |
| *Median (Q1, Q3)* | 83.00 (79.00, 88.00) | 85.00 (80.00, 90.00) | 68.00 (64.00, 71.00) | 69.00 (65.00, 72.00) | 67.00 (64.00, 71.00) | 69.00 (65.00, 72.00) | 67.00 (64.00, 70.00) | 69.00 (65.00, 72.00) | 68.00 (64.00, 72.00) | 69.00 (66.00, 72.00) |
| **Sex, n (%)** |  |  |  |  |  |  |  |  |  |  |
| *Female* | 308 (64.6) | 37,733 (53.0) | 188 (55.6) | 11,616 (45.3) | 116 (58.9) | 6,569 (48.7) | 27 (41.5) | 523 (41.0) | 104 (51.0) | 7,125 (42.2) |
| **Ethnicity, n (%)** |  |  |  |  |  |  |  |  |  |  |
| *White* | 426 (89.3) | 65,227 (91.5) | 298 (88.2) | 22,940 (89.5) | 172 (87.3) | 12,277 (91.0) | 56 (86.2) | 1,084 (84.9) | 182 (89.2) | 15,122 (89.6) |
| *Black* | 11 (2.3) | 898 (1.3) | 5 (1.5) | 345 (1.3) | 3 (1.5) | 139 (1.0) | <5 | 32 (2.5) | <5 | 220 (1.3) |
| *Asian* | 26 (5.5) | 2,185 (3.1) | 21 (6.2) | 1,138 (4.4) | 16 (8.1) | 524 (3.9) | <5 | 96 (7.5) | 12 (5.9) | 788 (4.7) |
| *Mixed* | <5 | 54 (0.1) | <5 | 39 (0.2) | <5 | 21 (0.2) | 0 (0.0) | 9 (0.7) | <5 | 29 (0.2) |
| *Other* | <5 | 240 (0.3) | <5 | 110 (0.4) | 0 (0.0) | 49 (0.4) | <5 | 9 (0.7) | <5 | 74 (0.4) |
| *Unknown* | 12 (2.5) | 2,648 (3.7) | 12 (3.6) | 1,073 (4.2) | 5 (2.5) | 480 (3.6) | <5 | 47 (3.7) | 6 (2.9) | 643 (3.8) |
| **Socioeconomic Status (IMD 2019 Quintiles), n (%)** |  |  |  |  |  |  |  |  |  |  |
| *1st quintile (least deprived)* | 90 (18.9) | 14,538 (20.4) | 52 (15.4) | 4,157 (16.2) | 22 (11.2) | 1,845 (13.7) | 18 (27.7) | 260 (20.4) | 30 (14.7) | 2,616 (15.5) |
| *2nd quintile* | 105 (22.0) | 15,451 (21.7) | 63 (18.6) | 4,597 (17.9) | 35 (17.8) | 2,221 (16.5) | 13 (20.0) | 265 (20.8) | 35 (17.2) | 2,912 (17.3) |
| *3rd quintile* | 98 (20.5) | 13,965 (19.6) | 66 (19.5) | 4,945 (19.3) | 36 (18.3) | 2,447 (18.1) | 15 (23.1) | 250 (19.6) | 41 (20.1) | 3,261 (19.3) |
| *4th quintile* | 83 (17.4) | 13,562 (19.0) | 82 (24.3) | 5,453 (21.3) | 49 (24.9) | 3,051 (22.6) | 12 (18.5) | 269 (21.1) | 47 (23.0) | 3,662 (21.7) |
| *5th quintile (most deprived)* | 101 (21.2) | 13,640 (19.1) | 75 (22.2) | 6,470 (25.2) | 55 (27.9) | 3,915 (29.0) | 7 (10.8) | 233 (18.2) | 51 (25.0) | 4,406 (26.1) |
| *Unknown* | 0 (0.0) | 96 (0.1) | 0 (0.0) | 23 (0.1) | 0 (0.0) | 11 (0.1) | 0 (0.0) | 0 (0.0) | 0 (0.0) | 19 (0.1) |

| Care home residence, n (%) |  |  |  |  |  |  |  |  |  |  |
| --- | --- | --- | --- | --- | --- | --- | --- | --- | --- | --- |
| Yes | <5 | 320 (0.4) | <5 | 118 (0.5) | <5 | 44 (0.3) | 0 (0.0) | 0 (0.0) | <5 | 66 (0.4) |
| Index year, n (%) |  |  |  |  |  |  |  |  |  |  |
| 2014 | 6 (1.3) | 3,974 (5.6) | 11 (3.3) | 1,317 (5.1) | 7 (3.6) | 672 (5.0) | 6 (9.2) | 56 (4.4) | 10 (4.9) | 819 (4.9) |
| 2015 | 23 (4.8) | 14,169 (19.9) | 17 (5.0) | 4,939 (19.3) | 11 (5.6) | 2,633 (19.5) | 5 (7.7) | 234 (18.3) | 9 (4.4) | 3,212 (19.0) |
| 2016 | 53 (11.1) | 15,339 (21.5) | 55 (16.3) | 5,647 (22.0) | 34 (17.3) | 2,931 (21.7) | 8 (12.3) | 276 (21.6) | 31 (15.2) | 3,666 (21.7) |
| 2017 | 92 (19.3) | 16,717 (23.5) | 76 (22.5) | 5,896 (23.0) | 42 (21.3) | 3,132 (23.2) | 9 (13.8) | 303 (23.7) | 49 (24.0) | 3,948 (23.4) |
| 2018 | 184 (38.6) | 16,317 (22.9) | 130 (38.5) | 6,038 (23.5) | 77 (39.1) | 3,142 (23.3) | 27 (41.5) | 313 (24.5) | 77 (37.7) | 4,034 (23.9) |
| 2019 | 119 (24.9) | 4,736 (6.6) | 49 (14.5) | 1,808 (7.1) | 26 (13.2) | 980 (7.3) | 10 (15.4) | 95 (7.4) | 28 (13.7) | 1,197 (7.1) |

Note: N = number of episodes
Abbreviations: Q1: first quartile; Q3: third quartile; RSV: respiratory syncytial virus; SD: standard deviation

**Table 2.**
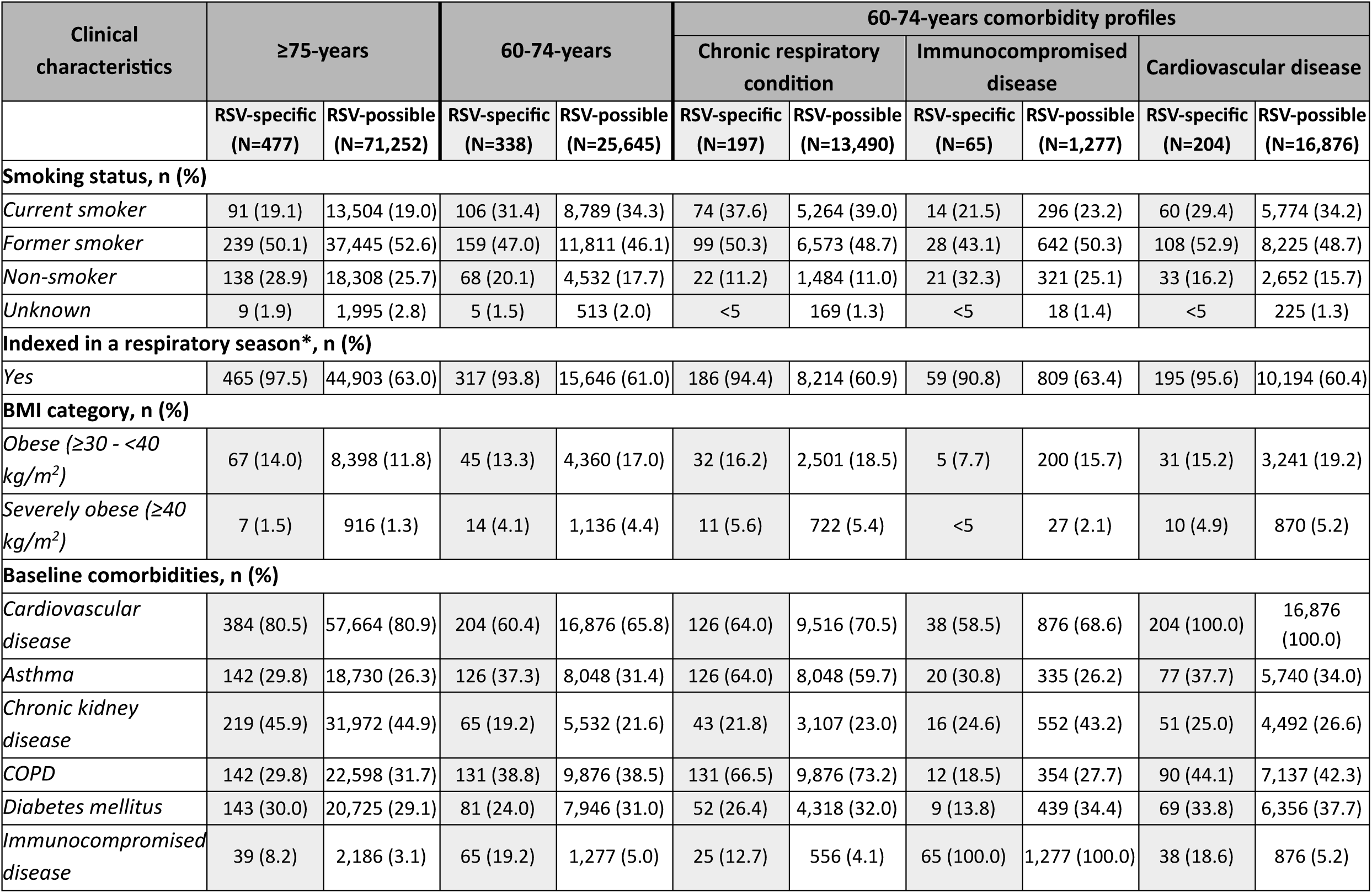
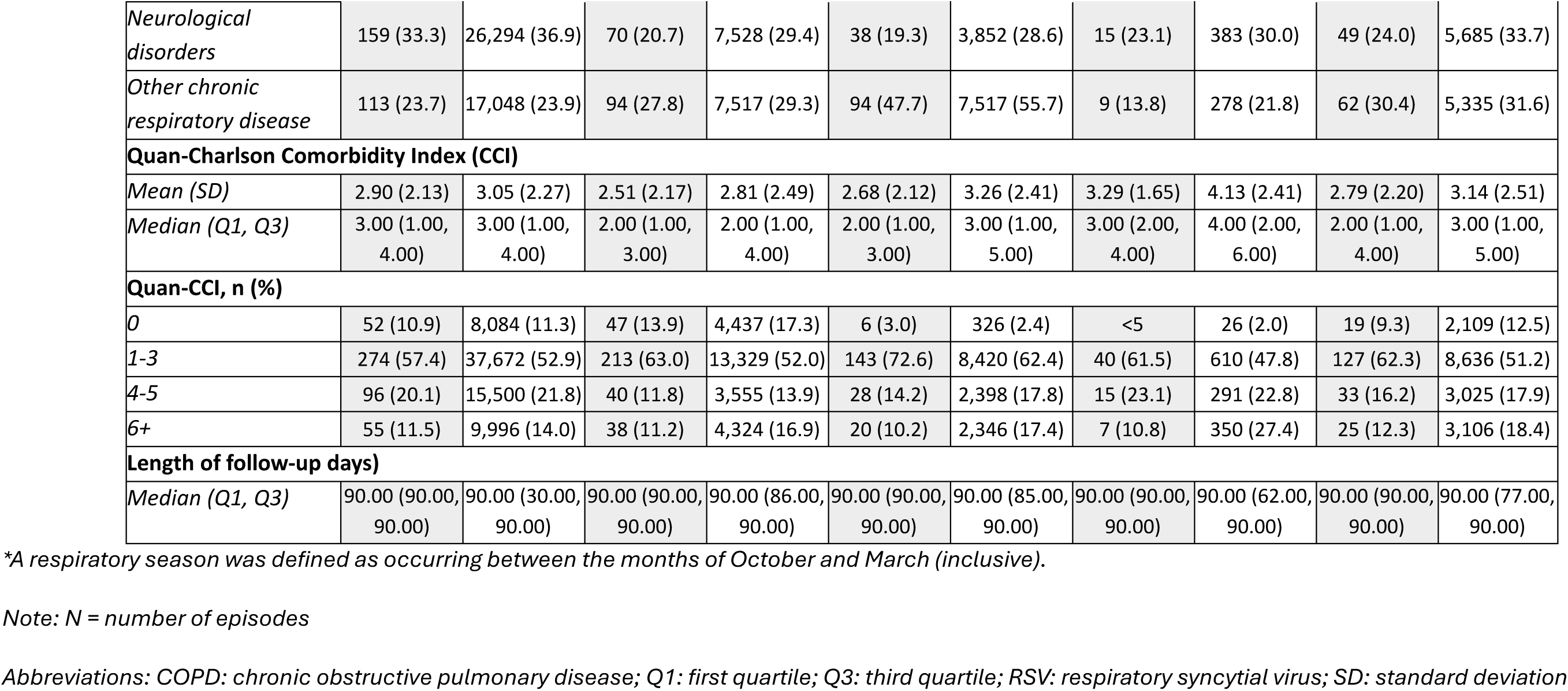
Baseline clinical characteristics among **RSV-specific and RSV-possible** episodes in those ≥75-years and 60-74-years of age, stratified by comorbidity profiles.

When descriptively comparing episodes in those 60-74-years of age to those ≥75-years of age, similarities were observed across most baseline characteristics. However, among episodes in patients aged ≥75-years, a higher Quan-CCI score was observed (mean Quan-CCI: 2.9 [SD: 2.1] vs. 2.5 [SD: 2.2]) compared to the younger age group.

#### RSV-possible case definition

Among 25,645 episodes (representing 23,672 patients) aged 60-74-years, a similar profile of baseline and clinical characteristics to the RSV-specific case definition episodes was observed; however the proportion of episodes indexed within a respiratory season was lower in the RSV-possible case definition compared to the RSV-specific episodes (possible: 61.0% [n=15,646] vs. specific: 93.8% [n=317]).

When descriptively comparing episodes under the RSV-possible case definition, to episodes in those under the RSV-specific case definition, similar profiles of baseline and clinical characteristics were observed in those aged ≥75-years.

### HCRU – Secondary care

#### RSV-specific case definition

The median (Q1-Q3) number of general inpatient admissions per-episode was 1.0 (1.0-2.0) across all age and comorbidity stratifications, except among those aged 60-74-years and immunocompromised (2.0 [1.0-3.0]) **(Table 3).** The median (IQR) general inpatient cumulative LoS per-episode among those aged ≥75-years was 10.0 (6.0-22.0) days which was higher than those aged 60-74-years: 9.0 [5.0-19.0] days. Among those aged 60-74-years stratified by specific comorbidities, LoS was comparable or higher than those aged ≥75-years, with the longest LoS reported in those immunocompromised (11.5 [7.0-26.0] days).

**Table 3.**
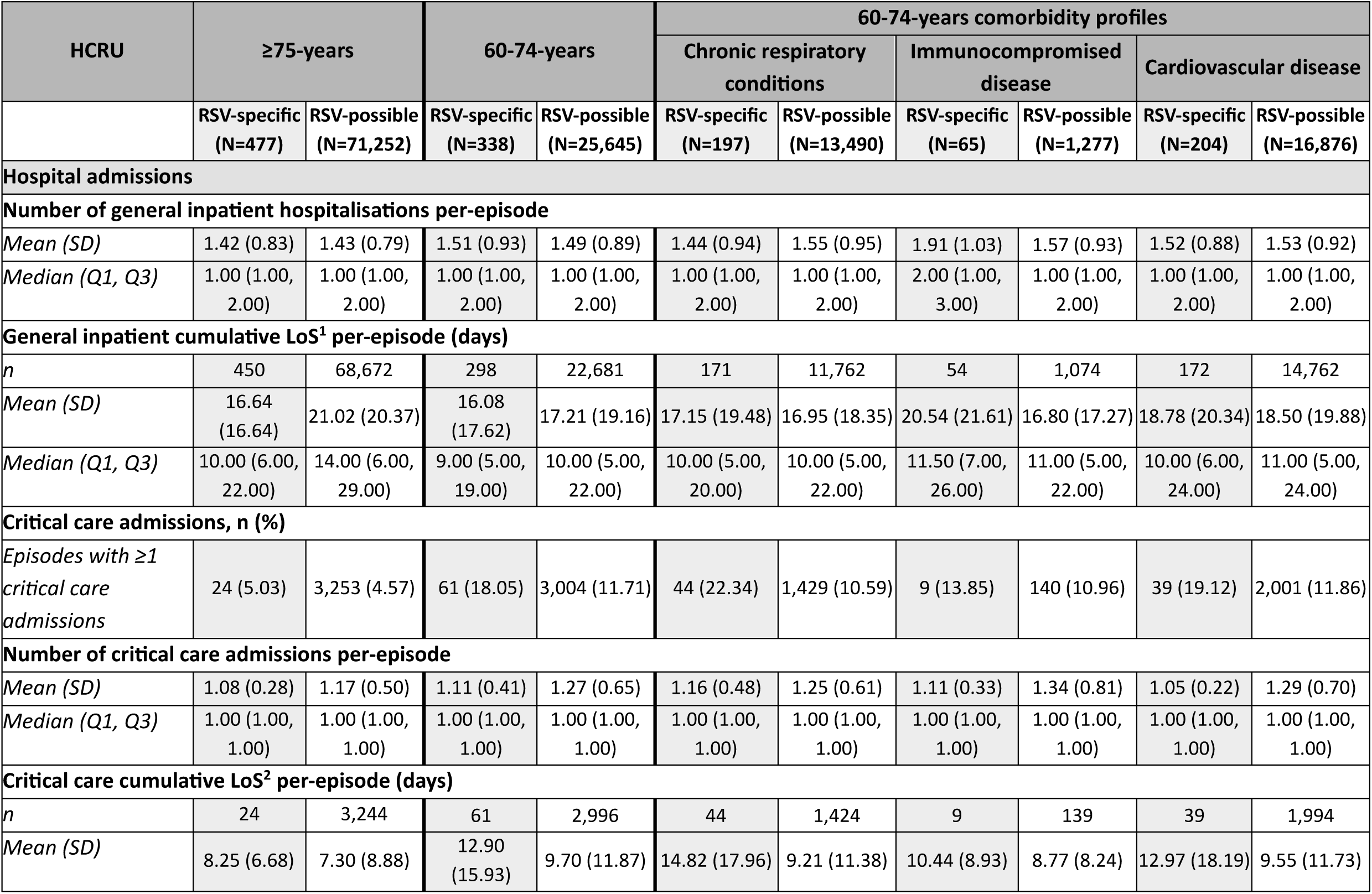
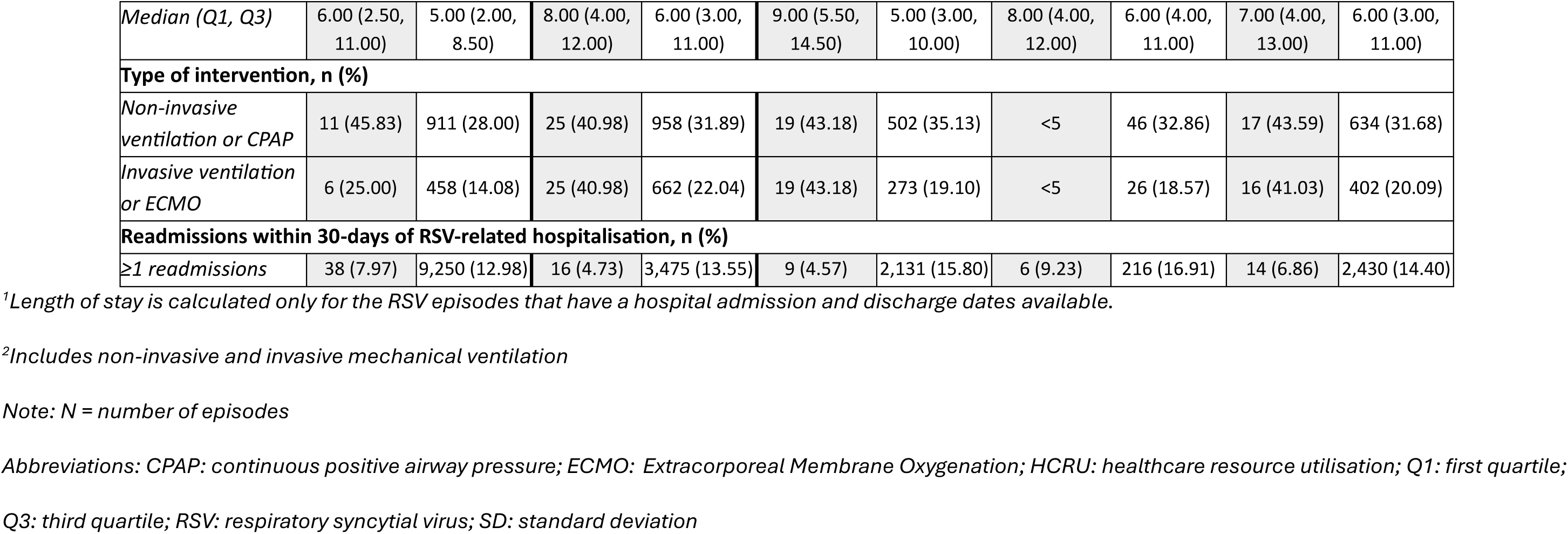
All-cause secondary care healthcare resource utilisation in the **G0-days after index** (RSV-episode) among **RSV-specific and RSV-possible** episodes in those ≥75-years of age and separately those aged 60-74-years, stratified by comorbidity profiles.

| HCRU | ≥75-years |  | 60-74-years |  | 60-74-years comorbidity profiles |  |  |  |  |  |
| --- | --- | --- | --- | --- | --- | --- | --- | --- | --- | --- |
|  |  |  |  |  | Chronic respiratory conditions |  | Immunocompromised disease |  | Cardiovascular disease |  |
|  | RSV-specific (N=477) | RSV-possible (N=71,252) | RSV-specific (N=338) | RSV-possible (N=25,645) | RSV-specific (N=197) | RSV-possible (N=13,490) | RSV-specific (N=65) | RSV-possible (N=1,277) | RSV-specific (N=204) | RSV-possible (N=16,876) |
| **Hospital admissions** |  |  |  |  |  |  |  |  |  |  |
| **Number of general inpatient hospitalisations per-episode** |  |  |  |  |  |  |  |  |  |  |
| *Mean (SD)* | 1.42 (0.83) | 1.43 (0.79) | 1.51 (0.93) | 1.49 (0.89) | 1.44 (0.94) | 1.55 (0.95) | 1.91 (1.03) | 1.57 (0.93) | 1.52 (0.88) | 1.53 (0.92) |
| *Median (Q1, Q3)* | 1.00 (1.00, 2.00) | 1.00 (1.00, 2.00) | 1.00 (1.00, 2.00) | 1.00 (1.00, 2.00) | 1.00 (1.00, 2.00) | 1.00 (1.00, 2.00) | 2.00 (1.00, 3.00) | 1.00 (1.00, 2.00) | 1.00 (1.00, 2.00) | 1.00 (1.00, 2.00) |
| **General inpatient cumulative LoS<sup>1</sup> per-episode (days)** |  |  |  |  |  |  |  |  |  |  |
| *n* | 450 | 68,672 | 298 | 22,681 | 171 | 11,762 | 54 | 1,074 | 172 | 14,762 |
| *Mean (SD)* | 16.64 (16.64) | 21.02 (20.37) | 16.08 (17.62) | 17.21 (19.16) | 17.15 (19.48) | 16.95 (18.35) | 20.54 (21.61) | 16.80 (17.27) | 18.78 (20.34) | 18.50 (19.88) |
| *Median (Q1, Q3)* | 10.00 (6.00, 22.00) | 14.00 (6.00, 29.00) | 9.00 (5.00, 19.00) | 10.00 (5.00, 22.00) | 10.00 (5.00, 20.00) | 10.00 (5.00, 22.00) | 11.50 (7.00, 26.00) | 11.00 (5.00, 22.00) | 10.00 (6.00, 24.00) | 11.00 (5.00, 24.00) |
| **Critical care admissions, n (%)** |  |  |  |  |  |  |  |  |  |  |
| *Episodes with ≥1 critical care admissions* | 24 (5.03) | 3,253 (4.57) | 61 (18.05) | 3,004 (11.71) | 44 (22.34) | 1,429 (10.59) | 9 (13.85) | 140 (10.96) | 39 (19.12) | 2,001 (11.86) |
| **Number of critical care admissions per-episode** |  |  |  |  |  |  |  |  |  |  |
| *Mean (SD)* | 1.08 (0.28) | 1.17 (0.50) | 1.11 (0.41) | 1.27 (0.65) | 1.16 (0.48) | 1.25 (0.61) | 1.11 (0.33) | 1.34 (0.81) | 1.05 (0.22) | 1.29 (0.70) |
| *Median (Q1, Q3)* | 1.00 (1.00, 1.00) | 1.00 (1.00, 1.00) | 1.00 (1.00, 1.00) | 1.00 (1.00, 1.00) | 1.00 (1.00, 1.00) | 1.00 (1.00, 1.00) | 1.00 (1.00, 1.00) | 1.00 (1.00, 1.00) | 1.00 (1.00, 1.00) | 1.00 (1.00, 1.00) |
| **Critical care cumulative LoS<sup>2</sup> per-episode (days)** |  |  |  |  |  |  |  |  |  |  |
| *n* | 24 | 3,244 | 61 | 2,996 | 44 | 1,424 | 9 | 139 | 39 | 1,994 |
| *Mean (SD)* | 8.25 (6.68) | 7.30 (8.88) | 12.90 (15.93) | 9.70 (11.87) | 14.82 (17.96) | 9.21 (11.38) | 10.44 (8.93) | 8.77 (8.24) | 12.97 (18.19) | 9.55 (11.73) |
| *Median (Q1, Q3)* | 6.00 (2.50, 11.00) | 5.00 (2.00, 8.50) | 8.00 (4.00, 12.00) | 6.00 (3.00, 11.00) | 9.00 (5.50, 14.50) | 5.00 (3.00, 10.00) | 8.00 (4.00, 12.00) | 6.00 (4.00, 11.00) | 7.00 (4.00, 13.00) | 6.00 (3.00, 11.00) |
| **Type of intervention, n (%)** |  |  |  |  |  |  |  |  |  |  |
| *Non-invasive ventilation or CPAP* | 11 (45.83) | 911 (28.00) | 25 (40.98) | 958 (31.89) | 19 (43.18) | 502 (35.13) | <5 | 46 (32.86) | 17 (43.59) | 634 (31.68) |
| *Invasive ventilation or ECMO* | 6 (25.00) | 458 (14.08) | 25 (40.98) | 662 (22.04) | 19 (43.18) | 273 (19.10) | <5 | 26 (18.57) | 16 (41.03) | 402 (20.09) |
| **Readmissions within 30-days of RSV-related hospitalisation, n (%)** |  |  |  |  |  |  |  |  |  |  |
| *≥1 readmissions* | 38 (7.97) | 9,250 (12.98) | 16 (4.73) | 3,475 (13.55) | 9 (4.57) | 2,131 (15.80) | 6 (9.23) | 216 (16.91) | 14 (6.86) | 2,430 (14.40) |

<sup>1</sup>Length of stay is calculated only for the RSV episodes that have a hospital admission and discharge dates available.
<sup>2</sup>Includes non-invasive and invasive mechanical ventilation
Note: N = number of episodes
Abbreviations: CPAP: continuous positive airway pressure; ECMO: Extracorporeal Membrane Oxygenation; HCRU: healthcare resource utilisation; Q1: first quartile;
Q3: third quartile; RSV: respiratory syncytial virus; SD: standard deviation

During an RSV episode, the proportion of 60-74-years olds admitted to critical care was higher than those aged ≥75-years (18.1% [n=61] vs 5.0% [n=24]); with median (IQR) cumulative LoS in critical care also longer at 8.0 (4.0-12.0) days vs 6.0 (2.5-11.0) days. The proportion requiring critical care and the median cumulative LoS in critical care was higher for all comorbidity stratifications in those aged 60-74-years, compared to those aged ≥75-years, with the greatest in those with chronic respiratory conditions (22.3% [n=44] and 9.0 [5.5-14.5] days, respectively). Furthermore, use of NIV/CPAP or invasive ventilation/ ECMO was also higher across all comorbidity stratifications when compared descriptively to episodes in those ≥75-years. The proportion of episodes with readmission to hospital within 30-days of an RSV-related hospitalisation was highest in those aged 60-74-years and immunocompromised (9.2% [n=6]).

#### RSV-possible case definition

Among those aged 60-74-years, similar patterns to the RSV-specific case definition were observed across secondary care HCRU. However, median (IQR) general inpatient cumulative LoS was slightly longer within the RSV-possible case definition (10.0 [5.0-22.0] days) versus the RSV-specific case definition (9.0 [6.0-22.0] days). The proportion of episodes that were admitted to critical care (possible: 11.7% [n=3,004]; specific: 18.1% [n=61]) and median (IQR) cumulative LoS (possible: 6.0 [3.0-11.0] days; specific: 8.0 [4.0-12.0] days) was higher within the RSV-specific case definition.

When descriptively comparing episodes under the RSV-possible case definition, to episodes under the RSV-specific case definition, similar trends in secondary care resource use were observed in patients aged ≥75-years.

Similar patterns to the RSV-specific case definition were observed when episodes in patients aged 60-74-years stratified by comorbidity profiles were descriptively compared to the ≥75-years group. Differences were observed in median (IQR) general inpatient cumulative LoS with episodes in those aged ≥75-years having longer stays (14.0 [6.0-29.0] days) compared descriptively to 60-74-years episodes stratified by comorbidity profiles. Additionally, the proportion of episodes requiring critical care and the cumulative LoS in critical care were higher and longer among the 60-74-years episodes stratified by comorbidity profiles compared to those aged ≥75-years.

### HCRU - Primary and Outpatient care

#### RSV-specific case definition

Approximately 80% of episodes had a primary care consultation during the 90-day RSV episode across all age and comorbidity stratifications, except among those aged 60-74-years and immunocompromised (61.5%) (**Table 4**). The mean number of medications prescribed in primary care ranged from 5.12 (SD: 5.57) in those aged 60-74-years and immunocompromised to 8.35 (SD: 6.92) in the same age group with chronic respiratory conditions. The most frequently prescribed medications were bronchodilators and antibiotics, with approximately 50% across all groups being prescribed antibiotics. Bronchodilators were more commonly prescribed in episodes of patients aged 60-74-years with chronic respiratory conditions (76.0%, n=111); while antibiotics were prescribed within 72.7% (n=24) of those immunocompromised in the same age group.

**Table 4.** All-cause primary care and outpatient healthcare resource utilisation in the **G0-days after index** (RSV-episode) among **RSV-specific and RSV-possible** episodes in those ≥75-years of age and separately those aged 60-74-years, stratified by comorbidity profiles.

| HCRU | ≥75-years |  | 60-74-years |  | 60-74-years comorbidity profiles |  |  |  |  |  |
| --- | --- | --- | --- | --- | --- | --- | --- | --- | --- | --- |
|  |  |  |  |  | Chronic respiratory conditions |  | Immunocompromised disease |  | Cardiovascular disease |  |
|  | RSV-specific (N=477) | RSV-possible (N=71,252) | RSV-specific (N=338) | RSV-possible (N=25,645) | RSV-specific (N=197) | RSV-possible (N=13,490) | RSV-specific (N=65) | RSV-possible (N=1,277) | RSV-specific (N=204) | RSV-possible (N=16,876) |
| **Primary care** |  |  |  |  |  |  |  |  |  |  |
| **Primary care consultations, n (%)** |  |  |  |  |  |  |  |  |  |  |
| *Episodes with ≥1 primary care consultations* | 388 (81.34) | 49,538 (69.53) | 267 (78.98) | 19,890 (77.56) | 159 (80.71) | 10,674 (79.13) | 40 (61.54) | 944 (73.92) | 159 (77.94) | 13,053 (77.35) |
| **Type of medications prescribed in primary care, n (%)** |  |  |  |  |  |  |  |  |  |  |
| *n* | 286 | 38,422 | 205 | 15,304 | 146 | 9,793 | 33 | 742 | 133 | 10,431 |
| *Antibiotics* | 140 (49.0) | 18,345 (47.7) | 113 (55.1) | 8,164 (53.3) | 78 (53.4) | 5,256 (53.7) | 24 (72.7) | 462 (62.3) | 74 (55.6) | 5,469 (52.4) |
| *Bronchodilators (long or short acting)* | 131 (45.8) | 15,489 (40.3) | 120 (58.5) | 8,427 (55.1) | 111 (76.0) | 7,431 (75.9) | 11 (33.3) | 260 (35.0) | 75 (56.4) | 5,693 (54.6) |
| *Inhaled corticosteroids* | 17 (5.9) | 1,704 (4.4) | 9 (4.4) | 797 (5.2) | 9 (6.2) | 679 (6.9) | 0 (0.0) | 26 (3.5) | 6 (4.5) | 510 (4.9) |
| *Oral steroids* | 62 (21.7) | 7,470 (19.4) | 59 (28.8) | 4,472 (29.2) | 49 (33.6) | 3,348 (34.2) | 0 (0.0) | 261 (35.2) | 38 (28.6) | 2,966 (28.4) |
| *Diuretics* | 140 (49.0) | 2,0876 (54.3) | 70 (34.1) | 6,326 (41.3) | 47 (32.2) | 3,830 (39.1) | 8 (24.2) | 269 (36.3) | 51 (38.3) | 4,917 (47.1) |
| **Number of medications prescribed in primary care** |  |  |  |  |  |  |  |  |  |  |
| *Mean (SD)* | 6.78 (7.70) | 6.26 (7.03) | 6.93 (6.52) | 6.77 (7.39) | 8.35 (6.92) | 8.51 (8.17) | 5.12 (5.57) | 5.15 (5.72) | 7.57 (7.05) | 7.24 (7.99) |
| **Outpatient care** |  |  |  |  |  |  |  |  |  |  |
| **RSV-related outpatient specialist encounters** |  |  |  |  |  |  |  |  |  |  |
| *Episodes with ≥1 RSV-related outpatient specialist encounters, n (%)* | 165 (34.59) | 23,273 (32.66) | 162 (47.93) | 12,588 (49.09) | 108 (54.82) | 7,177 (53.20) | 26 (40.00) | 684 (53.56) | 102 (50.00) | 8,650 (51.26) |
| *Mean (SD)* | 2.62 (3.43) | 2.27 (2.80) | 2.69 (2.90) | 2.70 (2.95) | 2.56 (2.82) | 2.68 (2.78) | 2.54 (2.21) | 3.20 (3.22) | 2.81 (2.65) | 2.75 (2.97) |
| *Median (Q1, Q3)* | 2.00 (1.00, 3.00) | 1.00 (1.00, 3.00) | 2.00 (1.00, 3.00) | 2.00 (1.00, 3.00) | 2.00 (1.00, 3.00) | 2.00 (1.00, 3.00) | 2.00 (1.00, 3.00) | 2.00 (1.00, 4.00) | 2.00 (1.00, 3.00) | 2.00 (1.00, 3.00) |

*Note: N = number of episodes*
*Abbreviations: HCRU: healthcare resource utilisation; Q1: first quartile; Q3: third quartile; RSV: respiratory syncytial virus; SD: standard deviation*

Nearly half of all episodes in patients aged 60-74-years attended an outpatient specialist consultation (47.9%, n=162), with closer to one-third amongst the ≥75-years group (34.6%, n=165). The median (IQR) outpatient consultations per episode was 2.0 (1.0-3.0) across all groups.

#### RSV-possible case definition

Similar patterns to the RSV-specific case definition were observed across primary care and outpatient consultations, including medications prescribed in primary care.

### Mortality

#### RSV-specific case definition

The all-cause case fatality rate for episodes in patients aged 60-74-years over a 90-day episode was 16.6% (n=56/338); while the RSV-related rate was 6.5% (n=22/338 [**Table 5**]), with similar proportions observed across comorbidity stratifications except chronic respiratory conditions (14.2% [n=28/197] and 5.1% [n=10/197], respectively). Both all-cause and RSV-related case fatality rates were higher amongst episodes in patients aged ≥75-years (all-cause: 19.3%; RSV-related: 10.3%).

**Table 5.** All-cause and RSV-related case fatality rates (90-days) among **RSV-specific and RSV-possible** episodes in those ≥75-years of age and separately those aged 60-74-years, stratified by comorbidity profiles.

|  | ≥75-years |  | 60-74-years |  | 60-74-years comorbidity profiles |  |  |  |  |  |
| --- | --- | --- | --- | --- | --- | --- | --- | --- | --- | --- |
|  |  |  |  |  | Chronic respiratory conditions |  | Immunocompromised disease |  | Cardiovascular disease |  |
|  | RSV-specific (N=477) | RSV-possible (N=71,252) | RSV-specific (N=338) | RSV-possible (N=25,645) | RSV-specific (N=197) | RSV-possible (N=13,490) | RSV-specific (N=65) | RSV-possible (N=1,277) | RSV-specific (N=204) | RSV-possible (N=16,876) |
| **All-cause case fatality (90-days)** |  |  |  |  |  |  |  |  |  |  |
| Total number of deaths, n (%) | 92 | 26,999 | 56 | 6,512 | 28 | 3,445 | 11 | 365 | 35 | 4,441 |
| Case fatality rate (CFR), % | 19.29% | 37.89% | 16.57% | 25.39% | 14.21% | 25.54% | 16.92% | 28.58% | 17.16% | 26.32% |
| **RSV-related case fatality (90-days)** |  |  |  |  |  |  |  |  |  |  |
| Total number of deaths, n (%) | 49 | 14,669 | 22 | 2,671 | 10 | 1,471 | <5 | 145 | 15 | 1,868 |
| Case fatality rate (CFR), % | 10.27% | 20.59% | 6.51% | 10.42% | 5.08% | 10.90% | <5 | 11.35% | 7.35% | 11.07% |

Note: N = number of episodes
Abbreviations: CI: confidence interval

#### RSV-possible case definition

All-cause and RSV-related case fatality rates were higher within the RSV-possible versus the RSV-specific case definition across all age and comorbidity stratifications. Similar trends were observed to the RSV-specific case definition, with both all-cause and RSV-related case fatality rates being highest among the ≥75-years group (all-cause: 37.9%; RSV-related: 20.6%).

## Discussion

To the best of our knowledge, this is the first study to quantify the HCRU associated with RSV by age and comorbidity profile among adults in England. We found that the burden of RSV in secondary care in adults aged 60-74-years across all comorbidities stratifications, is equivalent to or exceeds the burden of those aged ≥75-years. Notably, the proportion of episodes that required critical care admission was at least two times higher in those aged 60-74-years across comorbidities stratifications compared to those aged ≥75-years. Similar findings were observed in research conducted in Italy, whereby the proportion of patients hospitalised with RSV requiring critical care decreased as age increased; with higher proportions of admissions among patients aged ≥60-years deemed high-risk of severe illness due to comorbid conditions (31). We also found episodes in those aged 60-74-years with comorbid conditions had longer critical care LoS and greater use of mechanical ventilation. Selective critical care admission practices for frailer and multi-morbid patients may partly explain these findings; evidence suggests that some older patients, particularly those with comorbidities, poorer functional capacity and mental health, may be refused critical care as they could be deemed “too ill to benefit” following triage (32, 33).

The proportion of episodes requiring a primary care consultation was similar across all age groups and comorbidity stratifications. We observed over half of the episodes in those aged 60-74-years with comorbidities were prescribed antibiotics, even for the RSV-specific case definition. This finding mirrors both US and Italian-based studies, which found antibiotics to be administered to over half of adults with RSV aged ≥65-years and those with comorbid conditions that deemed them high-risk of severe illness (31, 34). Although regular prescriptions for other conditions could be attributable (reason for prescription is not recorded in the data), it may also reflect clinical practice whereby GPs initiate antibiotic therapy prior to PCR confirmed RSV infection. However, due to their lack of efficacy in treating viral infections and the increasing threat of antimicrobial resistance, a change in prescribing practices has been recommended by the UKHSA (35). Furthermore, as part of a holistic approach to antimicrobial resistance, high RSV vaccination rates could help reduce inappropriate antibiotic prescriptions and complement efforts to reduce antibiotic usage (36).

We observed higher all-cause and RSV-related case fatality rates among RSV episodes in those aged ≥75-years than among those aged 60-74-years with comorbidities, which is consistent with existing literature (31, 37) We also found stark differences between all-cause and RSV-related case fatality rates. The low rate in RSV-related mortality may be explained by the study observation period covering prior to the COVID-19 pandemic where established early detection viral testing strategies were introduced for older adults. Such limited testing may have resulted in RSV not being linked to a patient’s disease and so poorly recorded as cause of death.

Nearly half of RSV episodes were identified among patients in the most deprived socioeconomic quintiles, possibly reflecting a greater burden of RSV in more socioeconomically disadvantaged groups. This finding highlights the importance in equity in access to RSV prevention strategies available across the NHS schedule, as these patients may be less likely to have access to private healthcare. A low proportion of episodes were identified in care home residences, likely reflective of this data being poorly captured within CPRD rather than a low prevalence in this population. GPs were not required to document care home status prior to October 2020 and a patient’s residential address information in GP records can often be outdated (38).

We applied two distinct case definitions: RSV-specific and RSV-possible. Few differences were observed across between the two definitions when comparing baseline characteristics and HCRU between age groups and comorbidity profiles. A difference that was observed between these definitions was that the majority (>90%) of RSV-specific episodes were indexed during a respiratory virus season compared to ∼60% of RSV-possible episodes. This disparity is partly explained by seasonal patterns in both RSV activity and testing practices, whereby RSV diagnostic testing is preferentially performed during the peak respiratory season (October to March) (39). However, it should be noted that some out-of-season RSV-possible cases may not represent true RSV infections. The proportion of RSV-specific patients indexed did however increase towards the later-years of the indexing period, reflecting a growing practice of in-season PCR-testing.

Separately, we observed greater case fatality rates in RSV-possible episodes than RSV-specific episodes. These findings demonstrate the importance of exploring the burden of RSV across both case definitions. Whilst the RSV-specific case definition, provides an estimate of the burden in an RSV-confirmed population, the lack of routinely performed RSV testing means RSV illness and mortality is likely under recognised (40). Consequently, there is merit in supplementing the population using the RSV-possible case definition, particularly as it captures a larger sample of the population whilst noting lower specificity.

### Strengths

There are notable strengths to this study. The use of large and nationally representative datasets allowed for a well characterised RSV population across care settings and enhanced the generalisability to the general population in England. A study period encompassing five RSV respiratory seasons, enabled a more comprehensive assessment of disease burden over time, reducing the influence of season-specific variability. Finally, two complementary case definitions, with narrow (RSV-specific) and broader (RSV-possible) definitions allowed the establishment of upper and lower bounds for the burden of RSV in England.

### Study Limitations

Our study has several limitations. The RSV-specific and RSV-possible case definitions applied in this study have not been previously validated in the adult population; our findings may therefore underestimate or overestimate the RSV-specific and RSV-possible burden, respectively. We were unable to assess medications prescribed in the secondary care setting as CPRD Aurum only captures medications prescribed in primary care. Furthermore, we cannot distinguish between new and recurrent prescriptions, which means we cannot accurately attribute the medications prescribed during the RSV episode to the infection. Furthermore, the analysis in this study was purely descriptive and the analyses was not compared to a non-RSV control population, and therefore, the outcomes reported may not be RSV-related.

## Conclusions

Overall, our study highlights the clinical burden of RSV in adults aged 60-74-years with specific comorbidities, who may not be eligible for RSV vaccination in England, is equivalent to or exceeds the burden in those aged ≥75-years. These findings may inform health policy decision makers and highlight the need to prioritise the consideration of comorbidity groups that could benefit from RSV vaccination in the UK.

## Data Availability

All data produced in the present work are contained in the manuscript

## Acknowledgments

The authors thank Carmen Hockey for contributing to the conceptualisation of the work and Shuk-Li Collings for their critical review of the paper.

## Medical Writing, Editorial, and Other Assistance

N/A

## Authorship

Rebecca Butfield and Jack Said were involved with the conception and design of the work, and the interpretation of the data. Hannah Wright, Dexter Wiseman and Dheeraj Sethi were involved with the design of the work, and the interpretation of the data. Kiran Rai, Thomas Jennison, Jon Watkins and Isabel Jimenez were involved with the design of the work, and the acquisition and interpretation of the data. Antonia Geneidat was involved with the analysis of the data for the work. All authors critically reviewed the manuscript for important intellectual content and have participated sufficiently in the work to agree to be accountable for all aspects of the work in ensuring that questions related to the accuracy or integrity of any part of the work are appropriately investigated and resolved. All authors have given their final approval of the manuscript to be published.

## Ethics/Ethical Approval

This study used existing, fully de-identified data and the subject(s) cannot be identified, directly or through identifiers. Study results were in tabular form and aggregate analyses that omits subject identification. This study was approved by CPRD’s Research Data Governance (RDG) Committee (reference number: 24_004459) to access the data. This study also complied with all applicable laws regarding subject privacy, Declaration of Helsinki. No direct subject contact or primary collection of individual human subject data has occurred in this study.

## Data Availability

The data that support the findings of this study are available from the Clinical Practice Research Datalink (CPRD). Restrictions apply to the availability of these data, which were used under license for this study. Aggregated data are available from the authors with the permission of the CPRD.

